# DETECTION OF p53 AND SV40 IMMUNOSTAINING IN ARCHIVED TUMOR NEPHRECTOMY BLOCKS OF CHILDREN WITH NEPHROBLASTOMA IN KANO, NIGERIA

**DOI:** 10.1101/2025.10.20.25338183

**Authors:** Lofty John C. Anyanwu, Akinfenwa T. Atanda, Badamasi I Mohammed, Chioma J. Mba

## Abstract

**Background:** Immunohistochemical expression of p53 protein in tumor nephrectomy specimens of patients with nephroblastoma has been associated with an adverse clinical outcome. SV40 is a known tumorigenic virus associated with inflammatory kidney diseases. This study aimed to evaluate the role in clinical outcome of p53 and SV40 immunopositivity in formalin-fixed paraffin-embedded (FFPE) tissue blocks of children with nephroblastoma.

**Materials and methods:** This was a retrospective study of 24 nephroblastoma patients managed in a tertiary hospital. FFPE tumor tissue blocks from the patients were subjected to immunohistochemical staining for p53 and SV40, and their relationship to clinical outcome was analyzed using the methods of Kaplan – Meier. Statistical significance was set at p ≤0.05.

**Results:** The p53 immunopositivity rate in the study was 20.80% (5/24). None of the patient had a positive SV40 stain.

There was a significant association between p53 immunoreactive and gender (p=0.021). The study showed a statistically significant poorer overall survival (OS) for those who showed p53 immunopositivity (p=0.05).

**Conclusion:** The study provides preliminary evidence that p53 immunopositivity is associated with a poor survival. This may thus be used in the detection of patients with a poor prognostic tumor.

## Introduction

Nephroblastoma (also called Wilms tumor) is the most common malignant kidney tumor in children, with a prevalence of one in 10,000 children. It accounts for 80% of malignant tumors of the genitourinary system seen in children aged 15 years and below, and children less than 6 years of age account for 90% of cases of the disease. ^1,2,3^ About 75% of the children will be diagnosed before the age of 5 years. ^4,5^ The children commonly present with an abdominal mass (Fig. 1). The incidence of Nephroblastoma has been shown to very along ethnic groups and not geographic areas, with the negroid children (those of black African descent) having the highest rates, children of Asian descent having the lowest rates, while those of Caucasian descent have rates in between the two. ^4,6,7^

**Figure 1:**
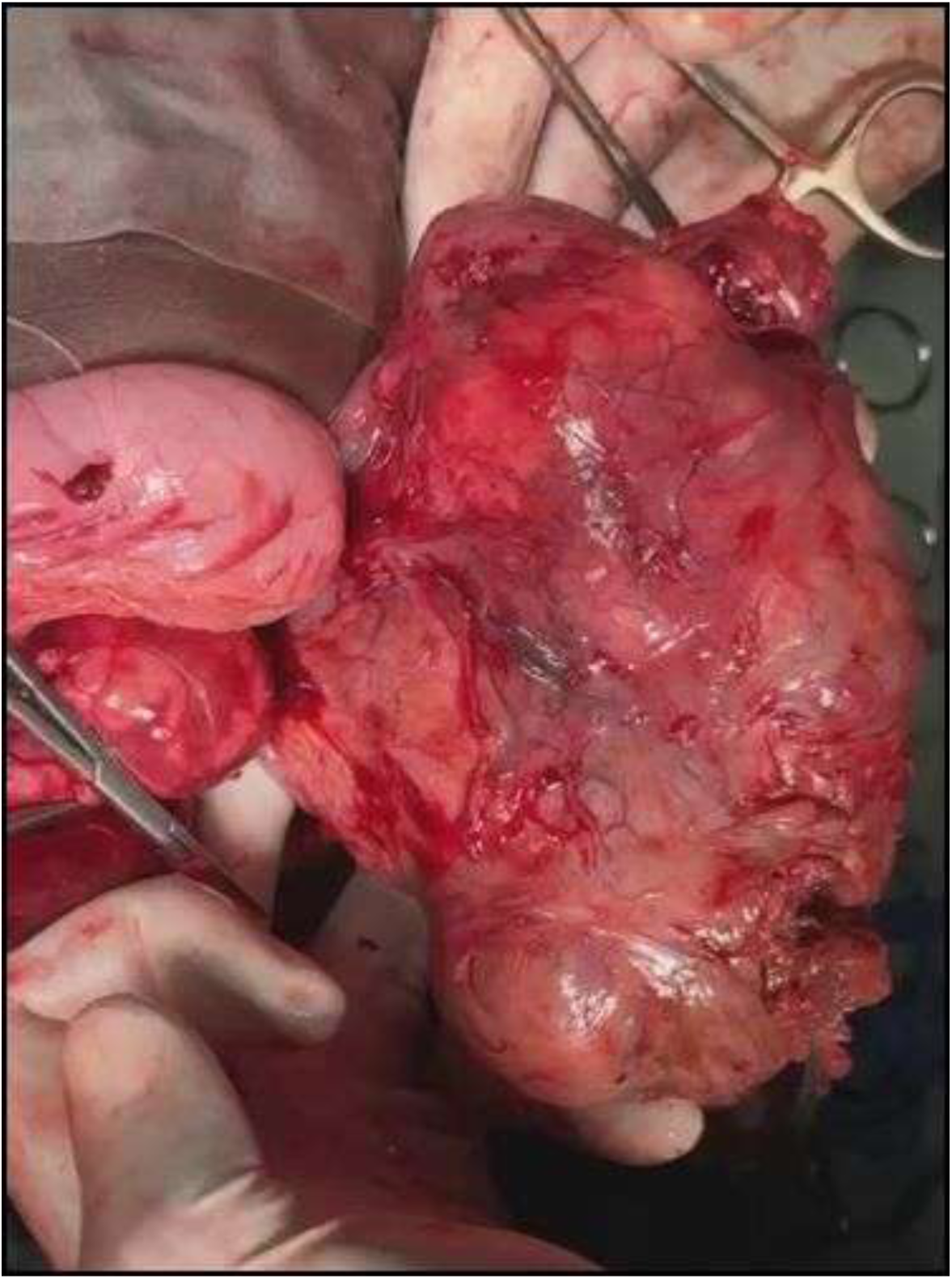
Intra-operative picture of a huge left Wilms tumor in a toddler.

The genetic basis for most cases of nephroblastoma is largely unknown, with only about 5% of patients with the disease having an identifiable genetic syndrome. ^6,8^ Commonly described genetic defects in nephroblastoma include mutations or deletions at locus 11p13 (WT1), which is present in 15% – 20% of sporadic cases, mutations or deletions at 11p15 (often called WT2) – which acts as a putative tumor suppressor gene. ^2,3,5^

In the treatment of nephroblastoma, a multimodal approach is commodity employed, and consists of surgery, chemotherapy and radiotherapy, depending on the stage and histology of the tumor. ^2,6,7^ For most developed countries, the 5 year overall survival of patients with nephroblastoma is about 90%, while for many sub-Saharan African countries, outcomes remain dismal, and 5 year overall survival as low as 25% have been reported. ^7,9,10,11^

Several studies have shown that p53 expression has a prognostic role in the outcome of patients with nephroblastoma, however the part played by p53 in the pathogenesis and progression of nephroblastoma is not fully understood.^12,13^ In the normal cell, the p53 tumor suppressor gene is located on chromosome 17p 13.1 and codes for p53 protein that acts as a multifunctional transcription factor involved in cell cycle checkpoint control, differentiation, senescence, apoptosis and DNA (deoxyribonucleic acid) replication and repair.^12,13,14^ Tumorigenesis and tumor progression are causally linked to p53 mutations, which may explain the irregular mitotic figures and increased DNA content seen in anaplastic nephroblastoma.^12,14^ Nuclear accumulation of abnormal protein is often (but not always) seen in tumors with p53 mutations.^15^ Immunodetectable p53 expression which signifies p53 gene mutation and abnormal accumulation of mutant p53 protein, is associated with both anaplasia and with an aggressive nephroblastoma with a poor prognosis.^3,14,15,16^

Given that cancer causing infections have been shown to be responsible for about 25% of cases of cancer in low-and middle-income countries,^17^ the potential role of oncogenic viruses as risk factors in the aetiopathogenesis of nephroblastoma, has not yet been fully elucidated. Mounting evidences suggest that the polyomavirus simian virus 40 (SV40) is a powerful DNA tumor virus known to be an emergent pathogen in humans. ^18,19^ SV40 Tag (large T antigen) causes abbreviations in the chromosomes of human cells which is thought to impede the functions of some genes involved in tumorigenesis such as DNA repair genes, oncogenes and tumor suppressor genes such as p53 and RB genes. ^19,20^ In humans, SV40 is associated with inflammatory kidney diseases, and certain cancers such as brain and bone tumors, lymphoma, and mesothelioma. ^18,19,20^ A recent report has shown SV40 immunohistochemical expression in formalin-fixed paraffin-embedded (FFPE) tissue blocks of renal cell carcinoma specimens.^21^ SV40 is believed to be transmitted in humans by both horizontal and vertical infection.^18,20,22^ This study aimed to evaluate the presence of immunohistochemical expression of p53 protein and SV40 stain – positivity in FFPE tissue blocks of children treated for nephroblastoma, and to assess their association with clinical outcome.

## Patients and Methods

### Study Design

This was a retrospective descriptive study. The study was conducted in the Department of Pathology and the Pediatric Surgery Unit of Department of Surgery of Aminu Kano Teaching Hospital (AKTH) in northwestern Nigeria. The hospital is a tertiary referral centre covering a catchment population of about 20 million people, and also attends to patients from neighboring countries like the Republic of Chad, and the Niger Republic.

### Patients’ Selection Process

Patients included in the study were those who were diagnosed and had surgery (tumor nephrectomy) for nephroblastoma between January 2014 and December 2023. Clinical data of the patients were retrospectively collected, as well as the retrieval of their formalin fixed paraffin – embedded (FFPE) tissue blocks. Only cases with accessible clinical data as well as available FFPE tissue blocks of good quality were included in the analysis. Patients with suboptimal tissue blocks and those with missing clinical data were excluded from the study. The study protocol was reviewed and approved by the research and ethics committee of AKTH Kano, Nigeria.

### Sampling Method

Using a convenient sampling method all cases with a histopathologically confirmed diagnosis of nephroblastoma who met the inclusion criteria were included in the study. All selected tissue blocks were reviewed by the pathologist (ATA) and classified histologically using the scheme of the International Society of Pediatric Oncologists (SIOP).^23^

### Immunohistochemistry

Tissue sections for immunohistochemistry were cut at 3 microns, floated onto APES-coated slides, blotted, heat fixed onto the slides at 70°C, dewaxed, rinsed in running tap water and distilled water. Antigen retrieval was done using citrate buffer in pressure cooker for 25 minutes. Endogenous peroxidase activity was blocked, rinsed in phosphate-buffered saline (PBS) and then treated with Ultra V protein block for 5 minutes to block for Non-specific background staining (NSB). The Ultra V was then drained from the slides. Diluted primary antibody dilution of 1 in 200 for p53 (DAKO clone DO-7), and 1 in 100 for SV40 virus (MAD clone PAb416) was added for 45 minutes, rinsed thoroughly with PBS/Tween20 and primary antibody enhancer was added for 10 minutes. Horse radish peroxidase polymer (HRP polymer) was applied for 10 minutes followed by rinsing in PBS/Tween20 and distilled water. This was followed by application of chromogenic substrate and immersion in 1% copper sulphate for 5 minutes each. The slides were then washed under running tap water, counterstained in Haematoxylin, dehydrated, cleaned and mounted.

### P53 Scoring

To ensure that density scores were reproducible and semiquantitative, a score of 0 was assigned if less than 5% of tumor cell nuclei expressed p53, 1 if 5% to 50% expressed p53 and 2 if more than 50% showed a positive p53 stain. For the assessment of intensity, a similar method was employed such that a score of 0 signified absent staining, 1 for intermediate intensity and 2 for strong intensity. Nuclear density and intensity scores were then combined to convert them to a third (composite) score. Those stains with an intensity and density score higher than 0 received a composite score of 1, while any other combination of staining density and intensity were scored as 0.^24^ Tonsil and colon specimens were used as positive and negative tissue controls for all runs of the immunohistochemical staining for p53.

### SV40 Stain scoring

SV40 stains were used in the detection of SV40 virus. Nuclelar staining without halo in the tumor cells were considered positive for SV40 virus. Confirmed SV40 virus negative and positive controls from renal allograft biopsied patients were used as controls. The staining intensity scores of target and control epithelial cells were graded from 0 to 3+, (0, no staining, or focal weak fine granular staining in <1%; 1+, weak fine granular staining in ≥1%; 2+, moderate granular staining; and 3+, strong granular staining).^25^

### Statistical Analysis

Data entry and analysis were done using the statistical program for social sciences (SPSS version 15.0 for windows). Continuous variables were presented as means +/- standard deviation, while frequencies and proportion were used for categorical variables. Associations between categorical variables were compared using the Chi-square test or the Fischer’s exact test where appropriate.

Overall survival (OS) was defined as time from diagnosis to death from any cause. Patients who were still alive were censored at the date of their last follow-up. Relapse free survival (RFS) was calculated from diagnosis to the date of the identification of new local or distant (metastatic) disease, or the date of patient’s last follow up. Estimates of RFS and OS were determined using the Kaplan-Meier method, and were compared between groups using the log-rank test. Cox proportional hazards regression analysis was performed to identify independent predictors of mortality (OS) and of disease progression or relapse (RFS). Patients’ follow-up was current through the December 31, 2024 data freeze. Statistical significance was set at p≤0.05 for all analysis.

## Results

### Patients’ characteristics

The clinical characteristics of the 24 patients who were included in the study is as summarized in Table 1. Their ages at presentation ranged from 1 year to 10 years, with a mean of 3.79years (SD 2.15years). Males made up 54.2% (13/24) of the participants, while females accounted for 45.8% (11/24), giving a male: female ratio of 1.18: 1. The intra-operative tumor stage for 17(70.8%) of the patients was stage III, while the remaining 7 patients (29.2%) had stage IV disease. Among the patients, 16 (66.7%) had a right sided tumor, while 8 (33.3%) had a left sided tumor. The median excised tumor weight was 950g (range 74g to 5000g). Mean length of the excised tumor was 16.38cm (SD, 3.95cm), with lengths ranging from 10cm to 25cm. Intra-operative tumor rupture occurred in nine (37.5%) of the patients. The most common histologic type was the stroma predominant nephroblastoma (8/24; 33.3%); followed by the mixed type nephroblastoma (7/24; 29.2%), blastema predominant type (6/24; 25.0%), epithelial predominant type (2/24; 8.3%), and the completely necrotic type (1/24; 4.2%). All the patients had pre-operative chemotherapy according to the SIOP-9 protocol.^26^ None of the histopathology reports in the patient records documented the presence of anaplasia.

**Table 1:**
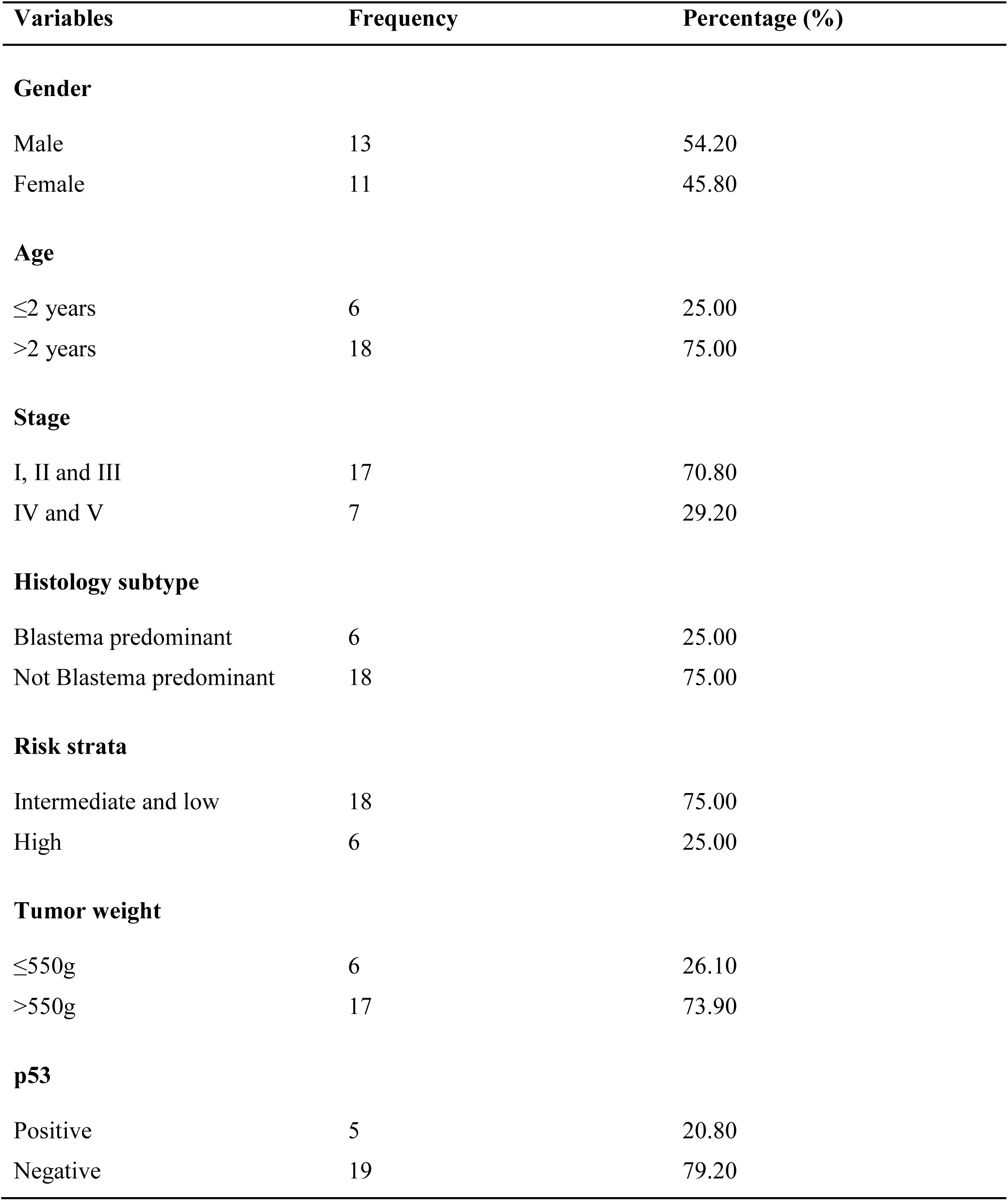
Characteristics of Study patients.

### Immunohistochemistry

SV40 stain positivity was not seen in all the 24 FFPE tumor blocks studied.

With regards to p53 immunohistochemical staining, five (20.80%) of the cases studied showed p53 immunohistochemical expression (Table 1; Fig. 2). The p53 scores are as shown in Table 2. p53 staining was seen in blastema, epithelia and stroma components of the tumors. There was no significant association between p53 expression and tumor stage (Table 3), or tumor weight (Table 4). There was also no significant association between p53 expression and age, histologic subtypes or risk stratification (Table 5). p53 expression was however significantly associated with gender (Table 5).

**Figure 2:**
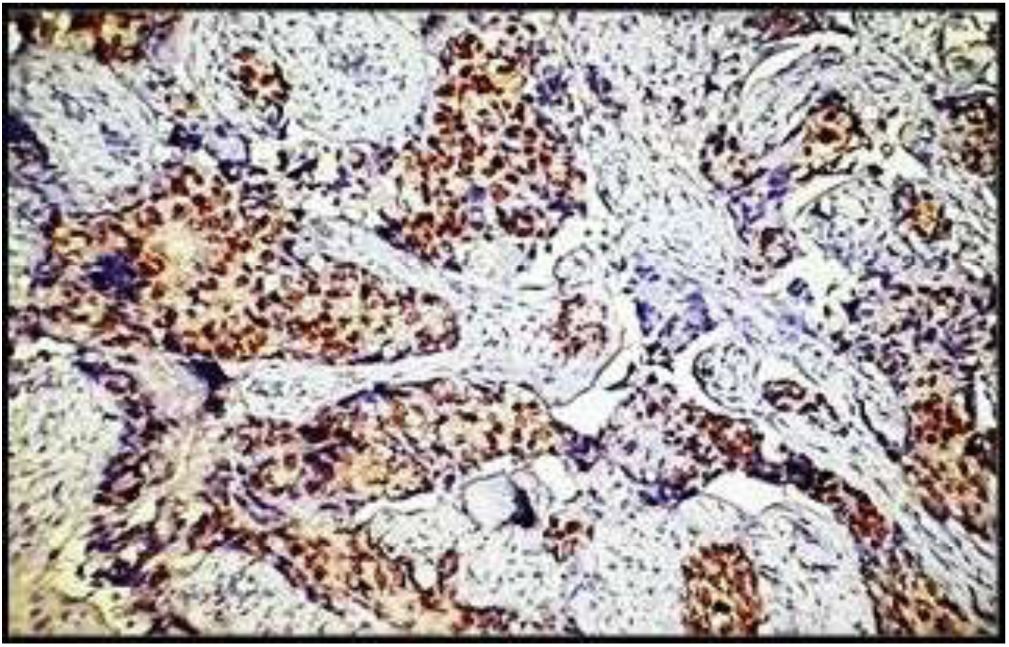
Neoplastic blastema cells showing nuclear p53 positivity surrounded by fibrous stroma. Anti-p53 Antibody x100.

**Table 2:** p53 Scores.

| **S/N** | **p53 Density** | **p53 Density Score** | **p53 Intensity Score** | **p53 Merged Score** |
| --- | --- | --- | --- | --- |
| **1** | 60 | 2 | 1 | 1 |
| **2** | 70 | 2 | 2 | 1 |
| **3** | 50 | 1 | 2 | 1 |
| **4** | 50 | 1 | 1 | 1 |
| **5** | 30 | 1 | 2 | 1 |

**Table 3:**
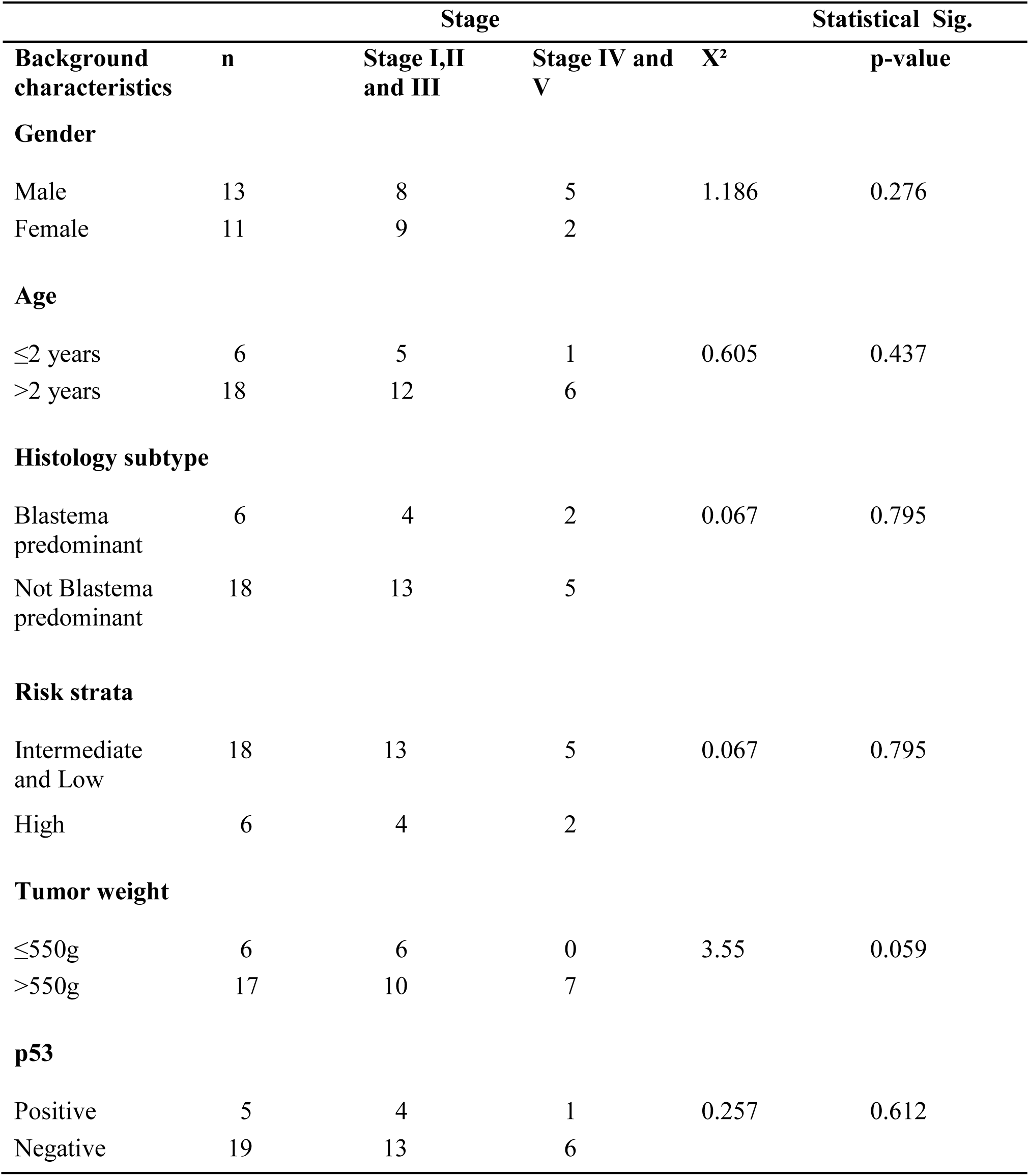
Tumor Stage in Relation to Patients’ Characteristics.

**Table 4:** Tumor Weight in Relation to Patients’ Characteristics.

|  |  | Tumor weight |  | Statistical significance |  |
| --- | --- | --- | --- | --- | --- |
| Background characteristics | n | ≤550g | >550g | X² | p-value |
| Gender |  |  |  |  |  |
| Male | 13 | 3 | 10 | 0.140 | 0.708 |
| Female | 10 | 3 | 7 |  |  |
| Age |  |  |  |  |  |
| ≤2 years | 6 | 4 | 2 | 6.933 | 0.008 |
| >2 years | 17 | 2 | 15 |  |  |
| Histologic subtype |  |  |  |  |  |
| Blastema predominant | 5 | 2 | 3 | 0.641 | 0.423 |
| Not Blastema predominant | 18 | 4 | 14 |  |  |
| Risk strata |  |  |  |  |  |
| Intermediate and low | 18 | 4 | 14 | 0.642 | 0.423 |
| High | 5 | 2 | 3 |  |  |
| p53 |  |  |  |  |  |
| Positive | 5 | 0 | 5 | 2.255 | 0.133 |
| Negative | 18 | 6 | 12 |  |  |

**Table 5:** p53 Staining in Relation to Patients’ Characteristics.

| Background characteristics | n | p53 staining |  | Statistical significance |  |
| --- | --- | --- | --- | --- | --- |
|  |  | Positive | Negative | X<sup>2</sup> | p-value |
| Gender |  |  |  |  |  |
| Male | 13 | 5 | 8 | 5.344 | 0.021 |
| Female | 11 | 0 | 11 |  |  |
| Age |  |  |  |  |  |
| ≤2 years | 6 | 0 | 6 | 2.105 | 0.147 |
| >2 years | 18 | 5 | 13 |  |  |
| Histologic subtype |  |  |  |  |  |
| Blastema predominant | 6 | 1 | 5 | 0.084 | 0.772 |
| Not Blastema predominant | 18 | 4 | 14 |  |  |
| Risk strata |  |  |  |  |  |
| Intermediate and Low | 18 | 4 | 14 | 0.084 | 0.772 |
| High | 6 | 1 | 5 |  |  |

### Outcome analysis

The median duration of follow-up after being discharged home following tumor nephrectomy among the patients was 8 months (range 0-83 months).

The two year relapse –free – survival (RFS) when all 24 patients were taken into account was 39%. However when the patients were classified according to stage of disease (Fig.3) RFS at 2years for stages I, II and III patients was 52%, while that for stages IV and V was 0%. The difference between the two was statistically significant (P<0.0001). Classification of the patients according to p53 immunoreactivity (Fig. 4), revealed that two-year RFS for patients who did not show p53 immunopositivity was 39%, while for those who were immunopositive for p53 it was 0%. The difference between the two was however not statistically significant (p = 0.560). The sites of relapse were commonly in the tumor bed and the liver.

**Figure 3:**
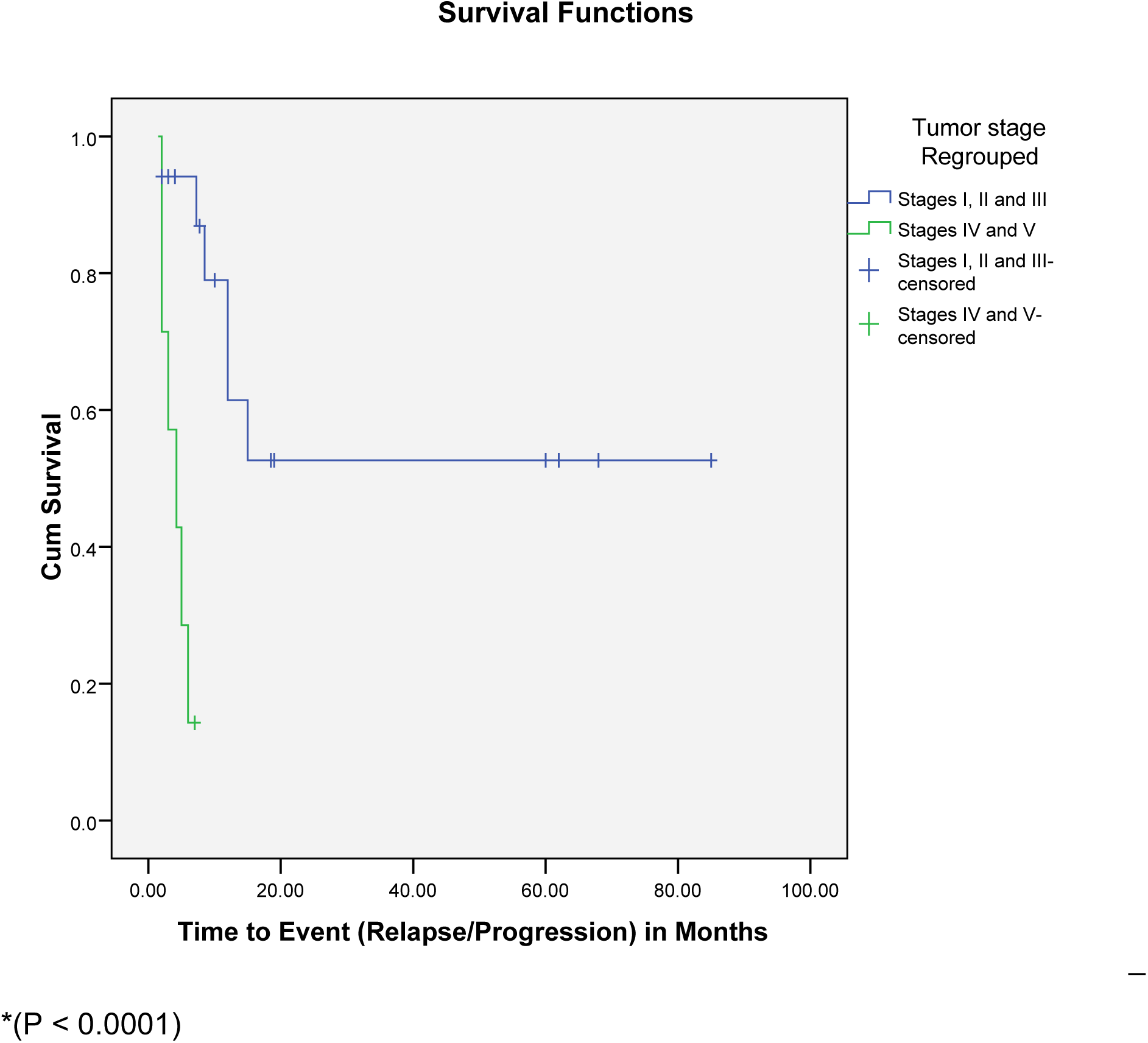
Two year RFS in patients with Wilms tumour according to stage.

**Figure 4:**
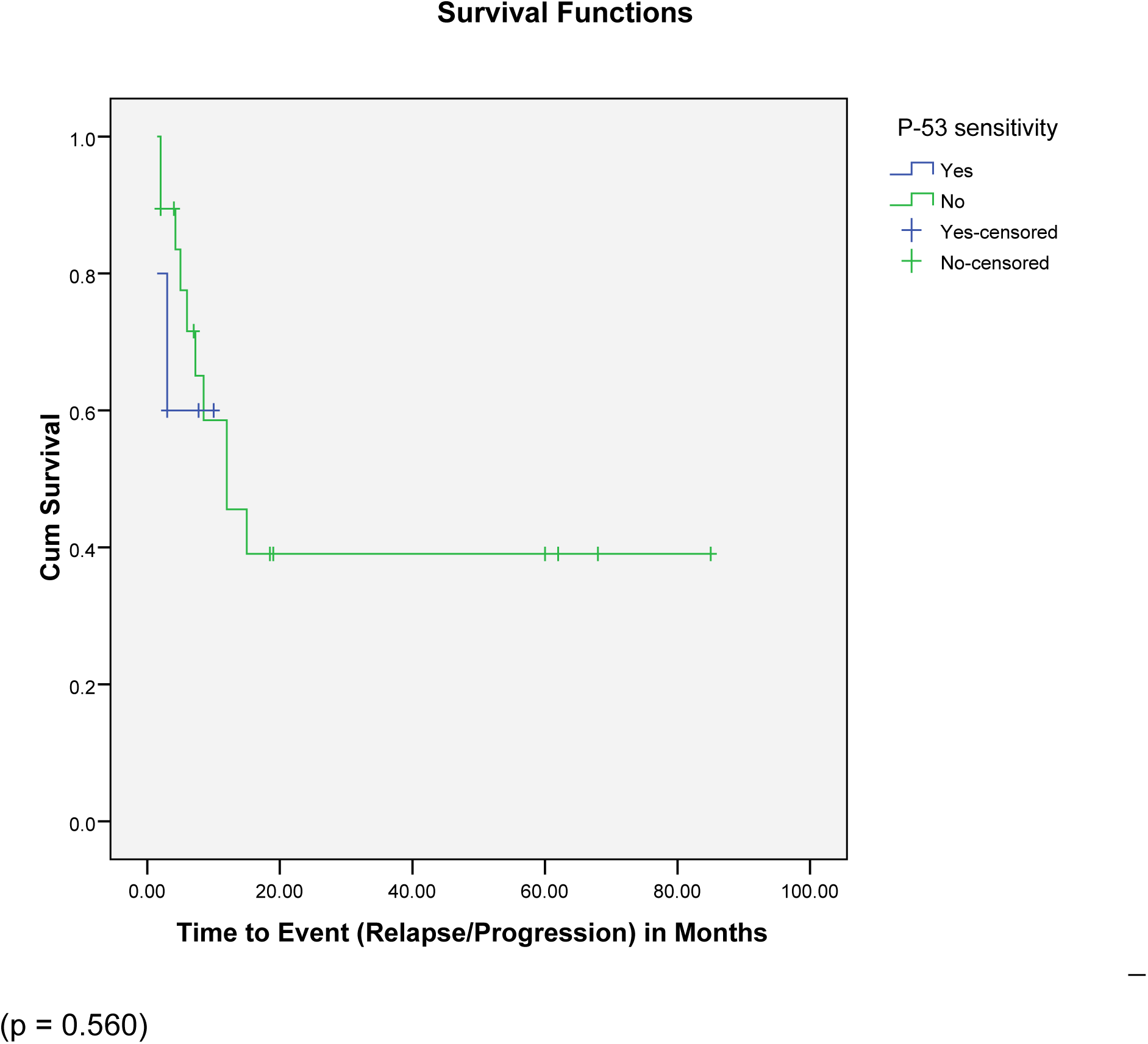
Two year RFS in patients with Wilms tumour according to p53 Immunopositivity.

Taking all patients into account overall survival (0S) at two years was 45%. Classification of the patients according to stage of disease (Fig. 5) showed that two year 0S for stages I, II and III patients was 53% while that for stage IV and V was 0%. The difference between the two was not statistically significant (p = 0.147). When the patients were classified according to p53 immunoreactivity (Fig. 6) however, the two-year 0S for those who did not show p53 immunopositivity was 52%, while for those who were immunopositive for P53 it was 0%. The difference between the two was statistically significant (p = 0.050).

**Figure 5:**
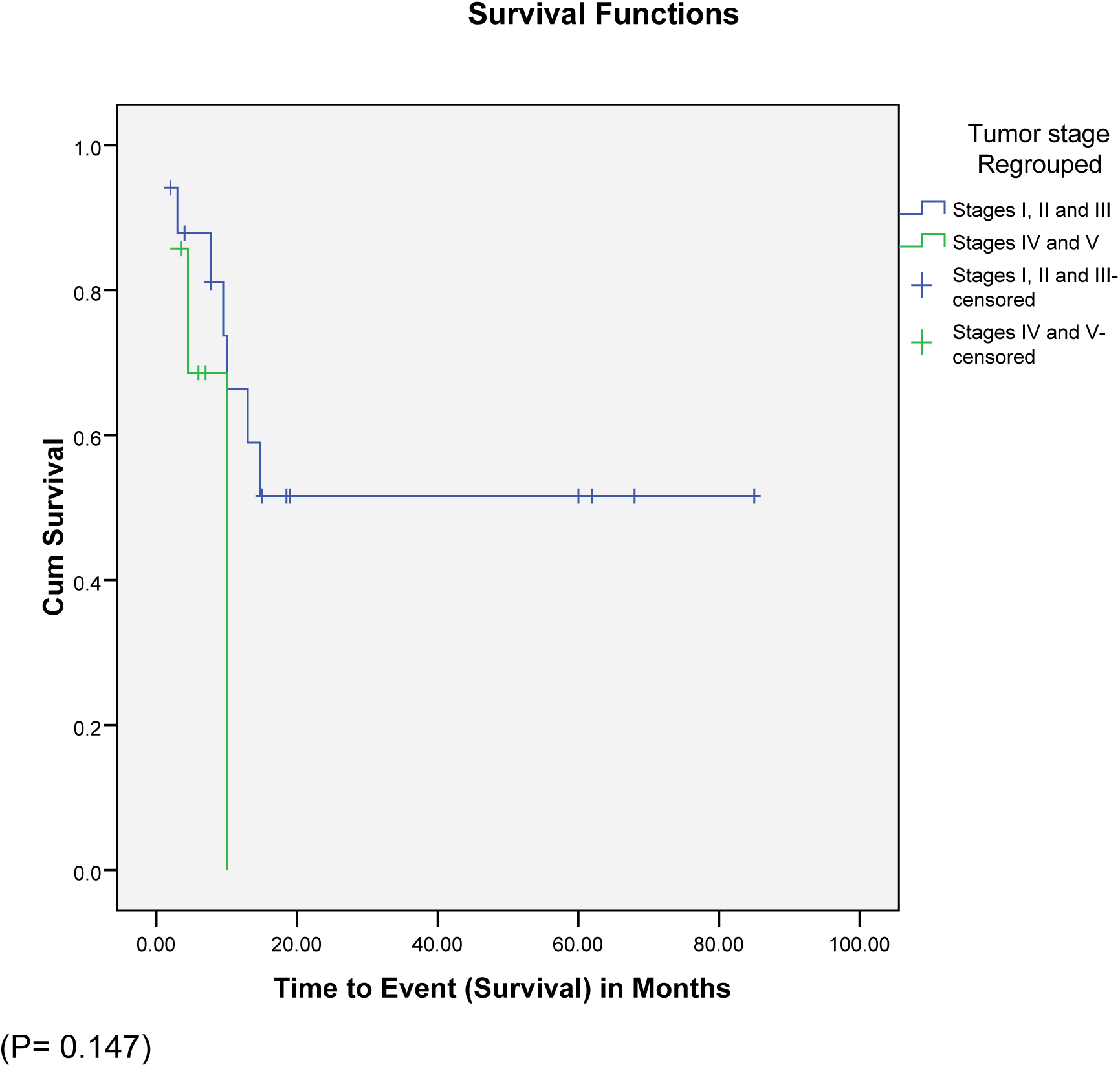
Two year OS in patients with Wilms tumour according to stage.

**Figure 6:**
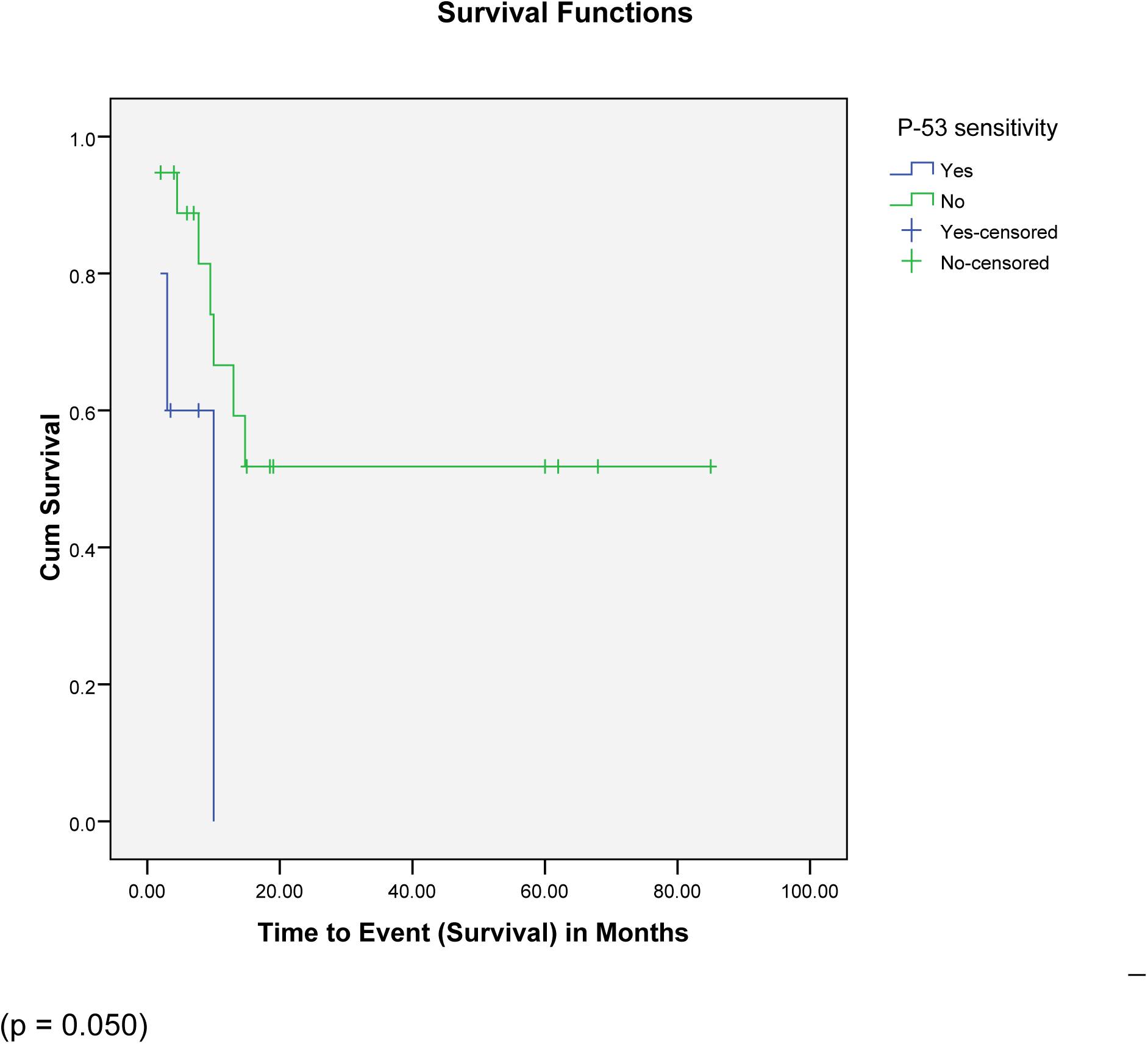
Two year OS in patients with Wilms tumour according to p53 Immunopositivity.

Tables 6 and 7, show the result of multivariate analysis using the Cox proportional hazards model to determine independent predictors of mortality (OS) and of disease relapse/progression (RFS). The model showed that disease stage was a significant predictor of disease relapse/progression, as patients with stage I, II and III disease, had only a 3.9% risk of relapse/progression compared to those with stages IV and V disease (Table 6). The model did not show any independent predictors of mortality among the patients (Table 7).

**Table 6:** Cox Regression Analysis of Independent Predictors of Disease Relapse/Progression.

| **Explanatory Variable** | **HR** | **p-value** | **95%CI**<br>**(For HR)** |  |
| --- | --- | --- | --- | --- |
| **Stage** | 0.039 | 0.005 | 0.004 | 0.366 |
| **Age** | 1.261 | 0.759 | 0.287 | 5.543 |
| **p53 sensitivity** | 2.958 | 0.260 | 0.449 | 19.496 |
| **Gender** | 1.219 | 0.770 | 0.324 | 4.582 |

*HR- Hazard ratio; CI – confidence interval*

**Table 7:** Cox Regression Analysis of Independent Predictors of Mortality.

| **Explanatory Variable** | **HR** | **p-value** | **95%CI**<br>**(For HR)** |  |
| --- | --- | --- | --- | --- |
| **Stage** | 0.224 | 0.098 | 0.038 | 1.317 |
| **Age** | 1.857 | 0.446 | 0.375 | 9.293 |
| **p53 sensitivity** | 6.099 | 0.066 | 0.884 | 42.058 |
| **Gender** | 0.954 | 0.955 | 0.191 | 4.772 |

*HR- Hazard ratio; CI – Confidence interval*

## Discussion

Nephroblastoma (Wilms’ tumor) is believed to occur most commonly in individuals of black African descent, being the second or third most frequently diagnosed childhood cancer in many parts of sub-Saharan Africa.^9^ Its peak age of occurrence is between 2 and 3 years of age, being uncommon in neonates.^3,4,5^ This is in keeping with the findings in this study where the mean age of the patients was 3.79years (SD 2.15years). Many have reported a slight female preponderance in those with Wilms’ tumor.^3, 4^ Our study however showed a male preponderance, which is at variance with similar studies from Egypt.^13, 27^

It has been shown that children presenting with Wilms’ tumor in sub-Saharan Africa, have a higher burden of disease which is characterized by an advanced disease stage, a higher tumor weight, and associated background malnutrition.^4,9,28,29^ The median excised tumor weight in this study was 950g (range 74g to 5000g), while the mean tumor length was 16.38cm (range 10cm to 25cm). An earlier report from Malawi showed that tumors in children with nephroblastoma (Wilms’ tumor) had a tumor diameter greater than 15cm in 70% of the case, and greater than 25cm in 28% of the cases, which were much larger than the mean tumor diameter of 9-10cm reported in the SIOP – 9 study in Europe.^30^ Given the documented difficulty associated with tumor nephrectomy for large Wilms’ tumors, many centers in sub-Saharan Africa (including ours), have adopted the use of the SIOP protocols, which incorporate obligatory pre-operative chemotherapy with the aim to improve the patients’ disease stage profile, and to reduce the risk of intra-operative tumor rupture.^10,11,28,30^ The intra-operative tumor rupture rate in this study was 37.5%. This is higher than the 21% reported by an earlier study from South Africa.^28^ Although we have adopted the SIOP – 9 protocol for the treatment of our Wilms’ tumor patients, we however do not give radiotherapy as it is not available in our center.

In this study 70.8% (17/24) of the patients had stage III disease, while 29.2% (7/24) had stage IV disease. This is in keeping with an earlier report from our center which showed that about 84% of patients with nephroblastoma had disease stages III and above on first presentation.^10,11^ This is however at variance with earlier reports from the National Wilms’ Tumor Studies 4 and 5, which showed that stage IV disease (metastatic nephroblastoma) occurred in only about 12.83% of all enrolled patients.^7,31^ Stage IV disease is known to carry a poor prognosis.^31^ This is in agreement with our findings (Table 6) which showed that having stage IV disease, was a significant predictor of disease relapse or progression.

Anaplastic histology in nephroblastoma is a feature of poor prognosis, with those with diffuse anaplasia having a significantly poorer prognosis than those with focal anaplasia.^2,8,32^ Although the frequency of anaplasia in the National Wilms’ Tumor Studies was between 5% - 13%, it is rare in the first 2 years of life, having a frequency of about 2%, but its frequency increases with age to about 13% in ages above 5 years.^2,32^ Anaplastic tumor cells have been shown to be neither generated by nor obliterated by chemotherapy and are believed to harbor p53 mutations and also to represent a more chemoresistant rather than a more aggressive tumor cell line.^2,23^ Anaplasia was not reported in the records of the patients enrolled in this study. This may be due to our small sample size. Detection of anaplasia will require a meticulous examination under the microscope of multiple sections of the tumor taken from each centimeter of maximal tumor diameter.^23,27^ Many authors have shown that p53 detection by immunostaining is strongly associated with the presence of anaplasia in Wilms’ tumors.^13,14,33^ Given the difficulties associated with the determination of the presence or absence of anaplasia in Wilms’ tumor specimens, it has been suggested that immunohistochemical detection of p53 can be used as a surrogate marker for anaplasia in tumor nephrectomy specimens.^14,33^ In this study, p53 immunostaining was positive in 20.80% of the cases (Table 1). This is less than the 60.3% reported by Salama and Kamel among Egyptian children.^27^It was however higher than the 13.0% reported from Uganda.^33^These findings may actually reflect regional differences. It may also reflect differences in the methods of scoring of p53 immunopositivity.^14^ This study showed that there was a higher risk of mortality in patients whose tumors showed p53 immunopositivity (Fig. 6).

In this study, none of the tissue blocks showed a positive SV40 immunostaining, this may be due to the problem of protein and nucleic acid degradation known to occur in formalin – fixed paraffin - embedded (FFPE) specimens.^18^ Although SV40 is a known tumorigenic virus in animal models, no comparable models to study its tumorigenesis in humans exist.^22^

One of the limitations of this study is its small sample size. Also the retrospective nature of the study meant that only data available in the medical records were used for analysis, and missing data could not be rectified.

## Conclusion

This study showed a significant elevation in the risk of mortality for patients whose tumors were immunopositive for p53 stain. A prospective study with a larger sample size will be needed to evaluate the real-time relationship between p53 immunopositivity and clinical outcomes in children with nephroblastoma in the region.

## Data Availability

Data is available on request from the corresponding author.

